# Effectiveness of BBV152 vaccine against SARS-CoV-2 infections, hospitalizations and deaths among healthcare workers in the setting of high delta variant transmission in New Delhi, India

**DOI:** 10.1101/2022.01.22.22269701

**Authors:** Sumit Malhotra, Mani Kalaivani, Rakesh Lodha, Sameer Bakhshi, Vijay Prakash Mathur, Pooja Gupta, Saurabh Kedia, Jeeva Sankar, Parmeshwar Kumar, Arvind Kumar, Vineet Ahuja, Subrata Sinha, Randeep Guleria

## Abstract

**Background:** Delta variant transmission resulted in surge of SARS CoV-2 cases in New Delhi, India during the early half of year 2021. Health Care Workers (HCWs) received vaccines on priority for prevention of infection. Real life effectiveness of BBV152 vaccine against severe disease including hospitalization and death was not known.

**Objective:** To estimate effectiveness of BBV152 vaccine among HCWs against SARS CoV-2 infection, hospitalization or death

**Design:** Observational study

**Setting:** a multi -speciality tertiary care public funded hospital in New Delhi, India.

**Participants:** 12,237 HCWs

**Interventions:** BBV152 vaccine (Covaxin, Bharat Biotech limited, Hyderabad, India); whole virion inactivated vaccine; two doses four weeks apart

**Measurements:** vaccine effectiveness after receipt of two doses of BBV152 protecting against any SARS CoV-2 infection, symptomatic infections or hospitalizations or deaths, and hospitalizations or deaths.

**Results:** The mean age of HCWs was 36(±11) years, 66% were men and 16% had comorbidity. After adjusting for potential covariates viz age, sex, health worker type category, body mass index, and comorbidity, the vaccine effectiveness (95% Confidence Interval) in fully vaccinated HCWs and ≥14 days elapsed after the receipt of second dose was 44% (37 to 51, p<0.001) against symptomatic infection, hospitalization or death due to SARS CoV-2, and 61% (37 to 76, p<0.001) against hospitalization or death, respectively.

**Conclusions:** BBV152 vaccine with complete two doses offer a modest response to SARS CoV-2 infection in real life situations against a backdrop of high delta variant community transmission. Efforts in maximizing receipt of full vaccines should be invested for HCWs, who are at higher occupational risk for infection.

## Introduction

The COVID-19 pandemic is continuing with multiple waves, which have been attributed to newer SARS-CoV-2 variants and waning immunity. Delta variant is the globally predominant circulating strain (1). India contributes to enormous high global case load with second largest cases reported (>34 million) so far in the world (2). The capital of India witnessed a recent surge of cases and experienced a second wave during months of March to May 2021, with a daily test positivity rate as high as 36% (3) and the surge in cases has been attributed to the delta variant (4).

India is also running one of the largest inoculation drives against COVID-19 with over 1 billion doses administered so far (5). Health Care Workers (HCWs) were the first priority group included for vaccination, starting from 16 January 2021. Two vaccines are being used-BBV152 (Covaxin, Bharat Biotech limited, Hyderabad) and ChAdOx1 (Covishield, Serum Institute of India, Pune). BBV152 is a whole virion inactivated vaccine with use of algel-IMDG (Imidazoquinoline molecule chemisorbed on alum) adjuvant to prime cell-mediated immune response (6). The complete series is with 2 doses administered, 4 weeks apart. The phase 3 trial of BBV152 vaccine conducted within India, reported efficacy of 78% and 93% against symptomatic and severe symptomatic infection (7). Recently, Covaxin has been included in the list for emergency use by World Health Organization (8). The efficacy of ChAdOx1 has been established outside Indian setting (9).

Real world effectiveness of COVID-19 vaccines is one of the important research priorities (10). Infection following COVID-19 vaccination is increasingly being reported after use of different vaccine types (11-13). Estimates of ChAdOx1 vaccine effectiveness are available in different settings, including for delta variant, both in India and other countries (14-16). There is no previous real time study reporting BBV152 vaccine effectiveness against hospitalization and death due to SARS-CoV-2 (17). We aimed to report effectiveness of BBV152 vaccine against SARS CoV-2 infections, hospitalizations and deaths among health care workers, in a setting of predominant transmission due to delta variant.

## Methods

### Study design

It was an observational study which collected data from different HCWs employed at the All-India Institute of Medical Sciences (AIIMS), New Delhi, India, a public-funded, teaching and multi-speciality tertiary care institute. The contact details of all HCWs (22,723) were obtained from administrative division, that maintains service records of all the HCWs within the Institute. The study period in this report was considered April 10 to June 24, 2021.

### Study participants

All employees (regular/contractual) and students at AIIMS were eligible to participate. Following categories of health care workers were invited to participate in the study: a) physicians (faculty, senior and junior residents and medical officers), b) nursing personnel, c) administrative and clerical staff, d) sanitation staff, e) security staff, f) social service supervisors, g) dietetics and related staff, h) laboratory personnel, i) technicians, j) research staff, k) scientists, l) health workers/ supervisors, m) paraclinical support staff, and n) students (medical, nursing, other undergraduates). Throughout the pandemic, AIIMS has been running a special employees’ health clinic for testing and management of COVID-19. Only BBV152 vaccine was offered at AIIMS and rolled out on 16 January 2021 onwards. HCWs who received Covishield were excluded from the current report.

### Data collection

All employees and students were contacted through e-mail, social media platforms like WhatsApp, and on telephone during May 12 to June 24, 2021. The data was collected either through a self-administered quality assurance measures in-built and web-based form (Google LLC, Mountain View, CA) or through telephone interview by trained personnel. The questionnaire included demographics (age/ gender/ type of HCW), co-morbidities, self-reported height/ weight, diagnosis of COVID-19 (yes/ no), history of vaccination (none/ single dose/ two doses and dates of receipt) and among those with positive diagnosis of COVID-19, information on the date of infection, number of episodes (single or more), mode of diagnosis [RT-PCR (Reverse Transcription Polymerase Chain Reaction), rapid antigen test (RAT), CBNAAT (Cartridge Based Nucleic Acid Amplification Test)], type of symptoms, severity including hospitalization details, and the outcome (recovery/ persistence of symptoms/ death). Additionally, reliable relatives of the deceased HCWs were interviewed telephonically to enquire about deaths during the study period, using the verbal autopsy method (18). All HCWs, who reported infection and 20% of not infected, were contacted again to validate the reported findings through vaccination cards and test reports.

### Definitions

1. **SARS CoV-2 infection** positivity detected by either molecular tests or antigen test (RT-PCR/ CBNAAT/ RAT) (11).
2. **Severity of disease** was graded as asymptomatic/ symptomatic; with details of symptomatic persons along with its severity (based on the World Health Organization (WHO) ordinal scale for clinical improvement), including hospitalization and death (19).
3. **Symptomatic SARS CoV-2 infection** Participant with any of the following symptoms was considered symptomatic-fever, rhinorrhoea, sore throat, cough, chest pain, wheezing, difficulty in breathing, shortness of breath, anosmia, dysgeusia, fatigue, myalgia, headache, abdominal pain, nausea, and diarrhoea.
4. **Comorbidity** considered were diabetes, hypertension, heart disease, chronic lung disease, chronic kidney disease, cancer, hypothyroidism and others.

### Statistical analyses

Data management and statistical analysis was done using Stata/SE 15.0 (StataCorp LLC, College Station, TX, USA). Categorical variables were presented as number (%) and continuous variables were presented as mean (SD)/ median (IQR).

Demographic and clinical variables such as age (<25, 25-44 and ≥45 years), sex (male and female), health care worker category (student/ administrative/clerical staff, faculty/scientist/research staff, nursing staff, junior/senior resident, paramedical/ support staff), Body Mass Index (BMI<18.5, 18.5-24.9 and ≥ 25 kg/m^2^) and comorbidity status (none and any comorbidity) with respect to SARS-CoV-2 infection and vaccination status was compared using Pearson’s chi-square test. The COVID-19 test positivity rate was very low during mid-January to beginning of April 2021. For assessing the vaccine effectiveness in this retrospective cohort study, we determined the exposure status as of 10 April 2021. On this date, the COVID-19 test positivity crossed 10% for the first time in Delhi (3). Also, this cut-off would provide for adequate numbers of subjects who would have received both doses of BBV152.

Based on the status of vaccine received, we had the following categories of subjects: **A**-unvaccinated throughout, also includes those who were unvaccinated as of 10 April 2021 and had the outcome (infection) before getting first dose of the vaccine; **B**-unvaccinated as of 10 April 2021, received dose 1 before end of the study, also includes those who received dose 1 of vaccine after 10 April 2021 and had the outcome (infection) before getting second dose of the vaccine; **C**-unvaccinated as of 10 April 2021 and received 2 doses before end of the study; **D**-partially vaccinated as of 10 April 2021, received second dose before end of the study; **E**-partially vaccinated throughout; and **F**-fully vaccinated as of 10 April 2021.

The vaccine effectiveness was estimated for three outcomes: 1) Any SARS-CoV-2 infection, 2) Symptomatic SARS-CoV-2 or Hospitalization or Death, 3) Hospitalization or Death due to SARS-CoV-2. The follow-up time was calculated from 10 April 2021 to date of diagnosis of infection for infected, from 10 April 2021 to the date of death for those who died due to SARS-CoV-2 infection and from 10 April to the date of interview for those who are not infected. To avoid bias, all the HCWs with deaths and prior infection of SARS-CoV-2 on or before 10 April 2021 were excluded from the analysis. The primary group of interest for determining the VE was ‘**F**’.

The probability of infection was estimated with respect to vaccination status using Kaplan-Meier curve and log rank test was used to compare the probability of infection among the groups. Next, we used the Poisson regression model between vaccination status and outcome controlling for demographic and clinical characteristics that could confound the association between vaccination and outcomes including age, sex, health care worker type, BMI and comorbidity status. From the estimated adjusted incidence rate ratio, we calculated the vaccine effectiveness (VE) by subtracting the incidence rate ratio from one (VE=1-incidence rate ratio). The p-value less than 0.05 was considered statistically significant.

## Results

### Study Flow and participants

This is shown in Figure 1. A total of 22,723 contact numbers of employees/ students were obtained from the administration and academic departments of AIIMS. These were contacted telephonically. Out of these, 7479 were excluded on account of ineligibility (1924), no response (3926), non -consent (672) and duplicate/ wrong entries (975). Thus, 15244 Health care workers (HCWs) finally participated in the study. Further, for vaccine effectiveness study, 2398 exclusions were done as 2380 HCWs got infected before 10 April 2021 and 18 HCWs died before 10 April 2021. HCWs (519) who received Covishield or did not know their vaccine type, were also excluded. Thus, in this current report data of 12,327 HCWs were analysed. The mean (SD) age of HCWs was 36 (11) years and 66% were men. The different categories of health care workers included were: paramedical/support staff (6871,55.7%), nursing staff (2410, 19.5%), students/administrative/clerical staff (1293,10.5%), and faculty/scientists/ research staff/physicians (1753,14.2%). The obesity (BMI >30Kg/m2) and any comorbidity was reported by 8% and 16% of HCWs respectively. The median follow-up of HCWs in this report was 48 days (Interquartile Range IQR 35 to 69 days).

**Figure 1.**
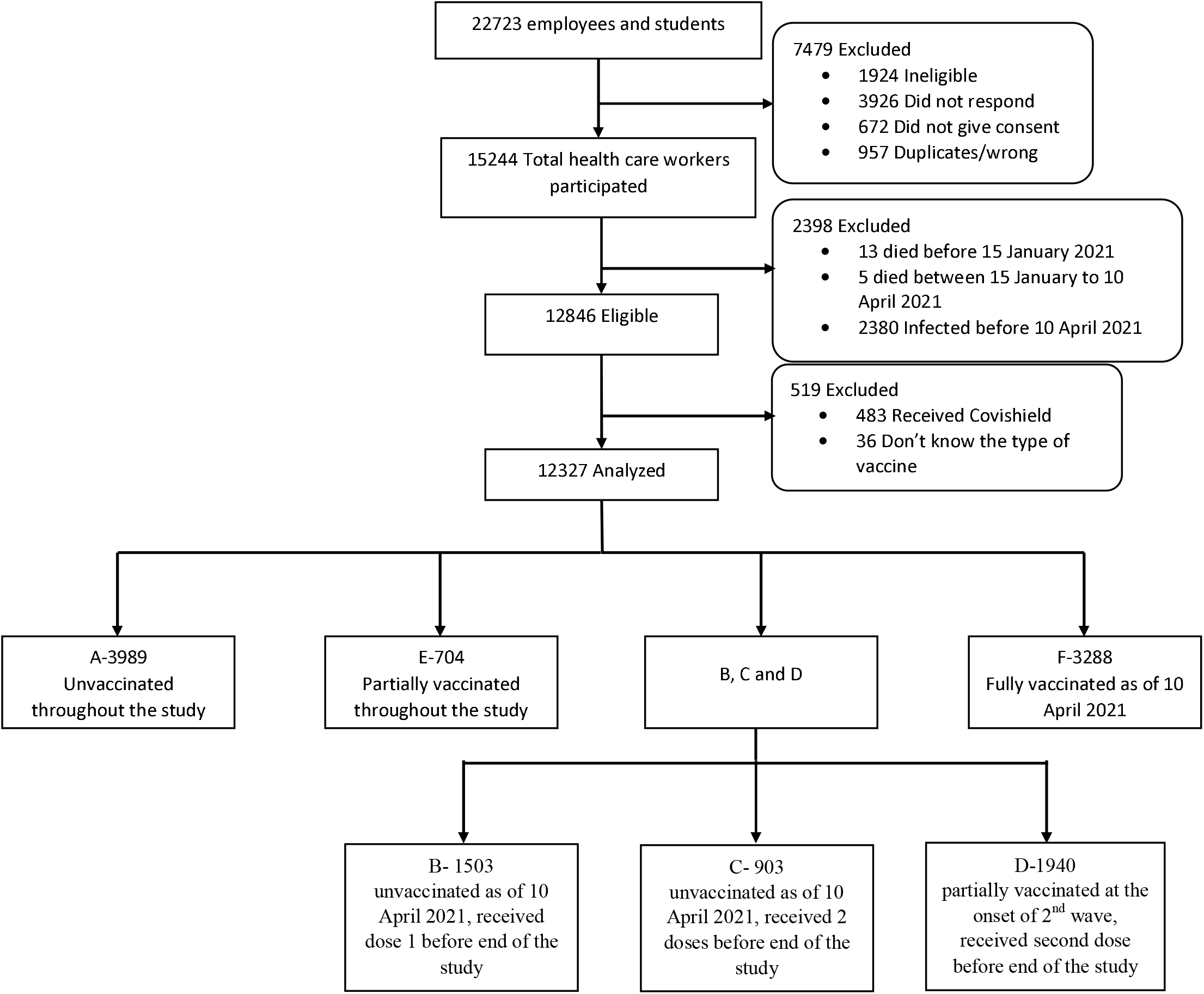
Study Flow Participants are health care workers who consented and either received at least one dose of BBV152 vaccine between 16 January 2021 and 24 June 2021 or did not receive vaccination. Analysis was carried out on 12237 participants to assess vaccine effectiveness among health care workers

### Vaccination exposures

Based on the vaccination status, HCWs were divided into six categories (**A** to **F**) considering the status on 10 April 2021. There were 3989 (32%) HCWs that were unvaccinated (**A**) and 3288 HCWs (27%) were fully vaccinated (**F**). Among fully vaccinated HCWs, there were 2208 HCWs with interval between second dose and 10 April ≥14 days. 704 HCWs were partially unvaccinated (**E**), throughout the study period. Remaining HCWs (4346) had different status of vaccination between 10 April 2021 to end of study period and distributed in following three - Category **B** comprised of HCWs (1503). Category **C** had 903 HCWs while Category **D** included 1940 HCWs.

### Baseline characteristics and SARS CoV-2 infection

The distribution of HCWs with respect to baseline demographic and clinical characteristics as per vaccination status is shown in Table 1 and there were significant differences observed. Among the study participants, 2484 (20%) reported to have any SARS CoV-2 infection. The incidence of SARS CoV-2 infection among unvaccinated (**A**) and fully vaccinated (**F**) HCWs was 26.5% (1056/3989) and 18.1% (595/3288), respectively. The median (IQR) interval between receipt of second dose and infection among category **F** HCWs was 47 (34,68) days.

**Table 1.** Details of diagnosed SARS-CoV-2 infections and vaccination status by demographic and clinical characteristics among health care workers.

|  | Total participants<br>(n=12327) | SARS-CoV-2 infection<br>(n=2484) | p-value<sup>a</sup> | Vaccination status |  |  |  |  |  | p-value<sup>a</sup> |
| --- | --- | --- | --- | --- | --- | --- | --- | --- | --- | --- |
|  |  |  |  | A<br>(n=3989) | B<br>(n=1503) | C<br>(n=903) | D<br>(n=1940) | E<br>(n=704) | F<br>(n=3288) |  |
| **Age in years** |  |  | <0.001 |  |  |  |  |  |  | <0.001 |
| < 25 | 1611 | 225 (14.0) |  | 618 (15.5) | 179 (11.9) | 94 (10.4) | 575 (14.2) | 121 (17.2) | 324 (9.9) |  |
| 25-44 | 7586 | 1676 (22.1) |  | 2539 (63.7) | 989 (65.8) | 526 (58.3) | 1117 (57.6) | 456 (64.8) | 1959 (59.6) |  |
| ≥45 | 3138 | 583 (18.6) |  | 832 (20.9) | 335 (22.3) | 282 (31.3) | 548 (28.3) | 127 (18.0) | 1005 (30.6) |  |
| **Gender** |  |  | <0.001 |  |  |  |  |  |  | <0.001 |
| Female | 4133 | 1164 (28.2) |  | 1418 (35.6) | 553 (36.8) | 278 (30.8) | 619 (31.9) | 283 (40.2) | 982 (29.9) |  |
| Male | 8194 | 1320 (16.1) |  | 2571 (64.4) | 950 (63.2) | 625 (69.2) | 1321 (68.1) | 421 (59.8) | 2306 (20.1) |  |
| **Health Care Worker Type** |  |  | <0.001 |  |  |  |  |  |  | <0.001 |
| Student/ Administrative/Clerical | 1293 | 203 (15.7) |  | 433 (10.9) | 125 (8.3) | 66 (7.3) | 250 (12.9) | 102 (14.5) | 317 (9.6) |  |
| Faculty/Scientist/Research | 893 | 245 (27.4) |  | 150 (3.8) | 106 (7.1) | 39 (4.3) | 131 (6.8) | 62 (8.8) | 405 (12.3) |  |
| Nursing | 2410 | 910 (37.8) |  | 867 (21.7) | 336 (22.4) | 164 (18.2) | 302 (15.6) | 158 (22.4) | 583 (17.7) |  |
| Junior/Senior Resident | 860 | 293 (34.1) |  | 109 (2.7) | 131 (8.7) | 37 (4.1) | 220 (11.3) | 107 (15.2) | 256 (7.8) |  |
| Paramedical/ Support Staff | 6871 | 833 (12.1) |  | 2430 (60.9) | 805 (53.6) | 597 (66.1) | 1037 (53.4) | 275 (39.1) | 1727 (52.4) |  |
| **Body Mass Index (Kg/m<sup>2</sup>)** |  |  | <0.001 |  |  |  |  |  |  | <0.001 |
| < 18.5 | 526 | 1291 (18.9) |  | 222 (5.6) | 64 (4.3) | 30 (3.3) | 82 (4.2) | 37 (5.3) | 91 (2.8) |  |
| 18.5-24.9 | 6832 | 64 (12.7) |  | 2231 (55.9) | 861 (57.3) | 497 (55.0) | 1077 (55.5) | 388 (55.1) | 1778 (54.1) |  |
|  |  |  |  |  | 469 (31.2) |  |  |  |  |  |
| 25.0-29.9 | 3927 | 893 (22.7) |  | 1197 (30.0) | 109 (7.3) | 295 (32.7) | 625 (22.2) | 223 (31.7) | 1118 (34.0) |  |
| ≥ 30 | 1042 | 233 (22.4) |  | 339 (8.5) |  | 81 (9.0) | 156 (8.0) | 56 (7.9) | 301 (9.1) |  |
| **Comorbidity** | 1928 | 492 (25.5) | <0.001 | 734 (18.4) | 207 (13.8) | 152 (16.8) | 278 (14.3) | 83 (11.8) | 474 (14.4) | <0.001 |

Data are number (%). Vaccination status (10 April 2021): A- unvaccinated throughout; also includes those who were unvaccinated as of 10 April 2021 and had the outcome (infection) before getting first dose of the vaccine; B-unvaccinated as on 10 April 2021, received dose 1 before end of the study; also includes those who received 1<sup>st</sup> dose of vaccine after 10 April 2021 and had the outcome (infection) before getting second dose of the vaccine; C- unvaccinated as on 10 April 2021, received 2 doses before end of the study; D- partially vaccinated as on 10 April 2021, received second dose before end of the study; E- partially vaccinated throughout and F- fully vaccinated as of 10 April 2021. <sup>a</sup> p-values (comparing the percentage of SARS-CoV-2 infections by demographic and clinical categories and comparing the percentage of vaccination status by these categories) calculated using Pearson's chi-square test.

**Table 2:** Person-days, incidence density of SARS-CoV-2 and vaccine effectiveness of BBV152 among health care workers.

| BBV152 immunization status | Person-days | SARS-CoV-2 infection events |  | Unadjusted |  | Adjusted |  |
| --- | --- | --- | --- | --- | --- | --- | --- |
|  |  | n | Incidence density/<br>10000 person-days<br>(95% CI) | VE<sup>a</sup> (95% CI) | p-value | VE<sup>a,b</sup><br>(95% CI) | p-value |
| **Any SARS-CoV-2 infection** |  |  |  |  |  |  |  |
| Unvaccinated (n= 3989)* | 167973 | 1056 | 62.9 (59.2, 66.8) | 1.0 |  | 1.0 |  |
| Fully vaccinated (n= 3288)** | 139020 | 595 | 42.8 (39.5, 46.4) | 0.32 (0.25, 0.38) | < 0.001 | 0.44 (0.37, 0.49) | < 0.001 |
| **Symptomatic SARS-CoV-2 or Hospitalization or Death** |  |  |  |  |  |  |  |
| Unvaccinated (n= 3989)* | 167973 | 966 | 57.5 (54.0, 61.3) | 1.0 |  | 1.0 |  |
| Fully vaccinated (n= 3288)** | 139020 | 378 | 38.5 (35.4, 41.9) | 0.33 (0.26, 0.40) | < 0.001 | 0.46 (0.39, 0.51) | < 0.001 |
| **Hospitalization or Death due to SARS-CoV-2** |  |  |  |  |  |  |  |
| Unvaccinated (n= 3989)* | 167973 | 73 | 4.3 (3.5, 5.5) | 1.0 |  | 1.0 |  |
| Fully vaccinated (n= 3288)** | 139020 | 31 | 2.2 (1.6, 3.2) | 0.49 (0.22, 0.34) | < 0.001 | 0.65 (0.46, 0.78) | < 0.001 |

Vaccination status (10 April 2021): \* unvaccinated throughout; also includes those who were unvaccinated as of 10 April 2021 and had the outcome (infection) before getting first dose of the vaccine (Category A); \*\* fully vaccinated as of 10 April 2021 (Category F). CI-Confidence Interval. Any SARS-CoV-2 includes death or hospitalization or symptomatic disease or asymptomatic infection. <sup>a</sup> Vaccine Effectiveness (VE) is estimated using the Poisson regression model, $VE=1-\text{incidence rate ratio}$ , <sup>b</sup> incidence rate ratios were adjusted for age, sex, health care worker category, body mass index and comorbidity

### Vaccine effectiveness

The total person days contributed by unvaccinated and fully vaccinated HCWs were 167973 and 139020, respectively. The incidence density for any SARS CoV-2 infection events among unvaccinated and fully vaccinated HCWs was 62.9 (95% CI 59.2 to 66.8) and 42.8 (95% CI 39.5 to 46.4) per 10000 person days, respectively. The adjusted vaccine effectiveness for preventing against any SARS CoV-2 infection was 43% (95% CI 36 to 48, p<0.001). The incidence density for symptomatic SARS CoV-2 infection or hospitalization or death among unvaccinated and fully vaccinated HCWs was 57.5 (95% CI 54.0 to 61.3) and 38.5 (95% CI 35.4 to 41.9) per 10000 person days, respectively. The adjusted vaccine effectiveness against same was 44% (95% CI 38 to 50, p<0.001). The incidence density for hospitalization or death due to SARS CoV-2 in unvaccinated and fully vaccinated HCWs was 4.3 (95% CI 3.5 to 5.5) and 2.2 (95% CI 1.6 to 3.2) per 10000 person days, respectively. The vaccine effectiveness against same was 65% (95% CI 46 to 78, p<0.001).

We also examined the vaccine effectiveness for fully vaccinated HCWs and segregating them as per the interval between the receipt of second dose and 10 April 2021. The vaccine effectiveness was almost similar for all the outcomes for HCWs with interval <14 days and ≥14 days (Table 3). The adjusted vaccine effectiveness in fully vaccinated HCWs ≥14 days for hospitalization or death due to SARS CoV-2 was 61% (95% CI 37 to 76, p<0.001). Varying significant effectiveness was found for HCWs in categories **B** to **D**, as shown in supplementary Table 1. Category **C** and **D** HCWs, both completing the full dose schedule before the end of study period, though starting the status on 10^th^ April 2021, with nil and partial vaccine status, had higher effectiveness of 99% and 80% respectively against both the outcomes-any SARS CoV-2 infection and symptomatic SARS CoV-2/ hospitalization/death. Vaccine effectiveness was not found among HCWs that were partially vaccinated throughout (category E of HCWs) as shown in Supplementary Table 1. The Kaplan -Meier curves and estimates are shown in Figure 2.

**Table 3:**
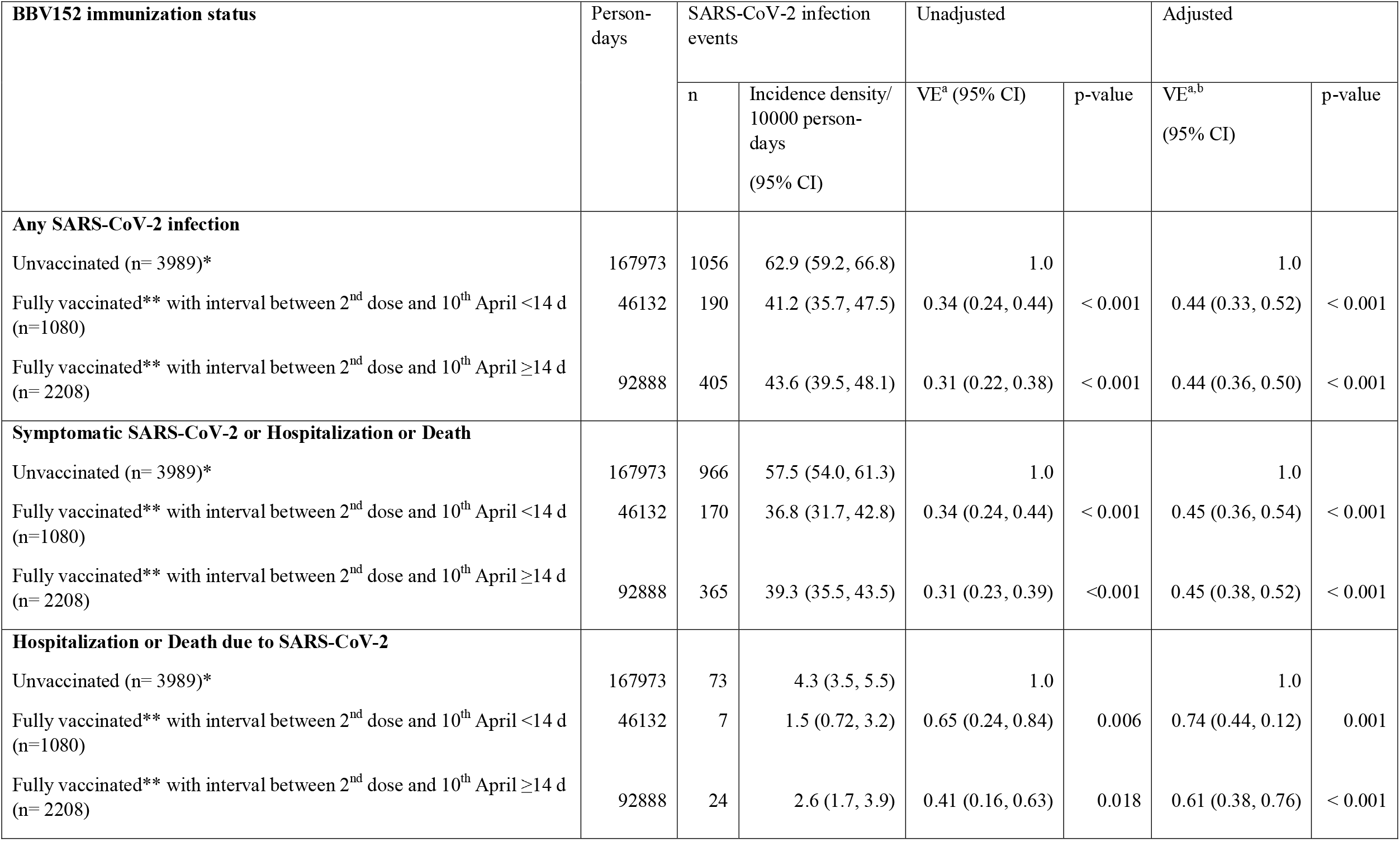
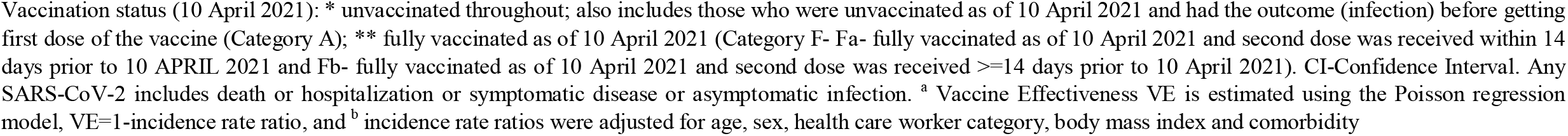
Person-days, incidence density of SARS-CoV-2 and vaccine effectiveness of BBV152 among health care workers.

| BBV152 immunization status | Person-days | SARS-CoV-2 infection events |  | Unadjusted |  | Adjusted |  |
| --- | --- | --- | --- | --- | --- | --- | --- |
|  |  | n | Incidence density/<br>10000 person-days<br>(95% CI) | VE<sup>a</sup> (95% CI) | p-value | VE<sup>a,b</sup><br>(95% CI) | p-value |
| **Any SARS-CoV-2 infection** |  |  |  |  |  |  |  |
| Unvaccinated (n= 3989)* | 167973 | 1056 | 62.9 (59.2, 66.8) | 1.0 |  | 1.0 |  |
| Fully vaccinated** with interval between 2<sup>nd</sup> dose and 10<sup>th</sup> April <14 d (n=1080) | 46132 | 190 | 41.2 (35.7, 47.5) | 0.34 (0.24, 0.44) | < 0.001 | 0.44 (0.33, 0.52) | < 0.001 |
| Fully vaccinated** with interval between 2<sup>nd</sup> dose and 10<sup>th</sup> April ≥14 d (n= 2208) | 92888 | 405 | 43.6 (39.5, 48.1) | 0.31 (0.22, 0.38) | < 0.001 | 0.44 (0.36, 0.50) | < 0.001 |
| **Symptomatic SARS-CoV-2 or Hospitalization or Death** |  |  |  |  |  |  |  |
| Unvaccinated (n= 3989)* | 167973 | 966 | 57.5 (54.0, 61.3) | 1.0 |  | 1.0 |  |
| Fully vaccinated** with interval between 2<sup>nd</sup> dose and 10<sup>th</sup> April <14 d (n=1080) | 46132 | 170 | 36.8 (31.7, 42.8) | 0.34 (0.24, 0.44) | < 0.001 | 0.45 (0.36, 0.54) | < 0.001 |
| Fully vaccinated** with interval between 2<sup>nd</sup> dose and 10<sup>th</sup> April ≥14 d (n= 2208) | 92888 | 365 | 39.3 (35.5, 43.5) | 0.31 (0.23, 0.39) | <0.001 | 0.45 (0.38, 0.52) | < 0.001 |
| **Hospitalization or Death due to SARS-CoV-2** |  |  |  |  |  |  |  |
| Unvaccinated (n= 3989)* | 167973 | 73 | 4.3 (3.5, 5.5) | 1.0 |  | 1.0 |  |
| Fully vaccinated** with interval between 2<sup>nd</sup> dose and 10<sup>th</sup> April <14 d (n=1080) | 46132 | 7 | 1.5 (0.72, 3.2) | 0.65 (0.24, 0.84) | 0.006 | 0.74 (0.44, 0.12) | 0.001 |
| Fully vaccinated** with interval between 2<sup>nd</sup> dose and 10<sup>th</sup> April ≥14 d (n= 2208) | 92888 | 24 | 2.6 (1.7, 3.9) | 0.41 (0.16, 0.63) | 0.018 | 0.61 (0.38, 0.76) | < 0.001 |

Vaccination status (10 April 2021): \* unvaccinated throughout; also includes those who were unvaccinated as of 10 April 2021 and had the outcome (infection) before getting first dose of the vaccine (Category A); \*\* fully vaccinated as of 10 April 2021 (Category F- Fa- fully vaccinated as of 10 April 2021 and second dose was received within 14 days prior to 10 APRIL 2021 and Fb- fully vaccinated as of 10 April 2021 and second dose was received $\geq 14$ days prior to 10 April 2021). CI-Confidence Interval. Any SARS-CoV-2 includes death or hospitalization or symptomatic disease or asymptomatic infection. <sup>a</sup> Vaccine Effectiveness VE is estimated using the Poisson regression model, $VE=1$ -incidence rate ratio, and <sup>b</sup> incidence rate ratios were adjusted for age, sex, health care worker category, body mass index and comorbidity

**Figure 2.**
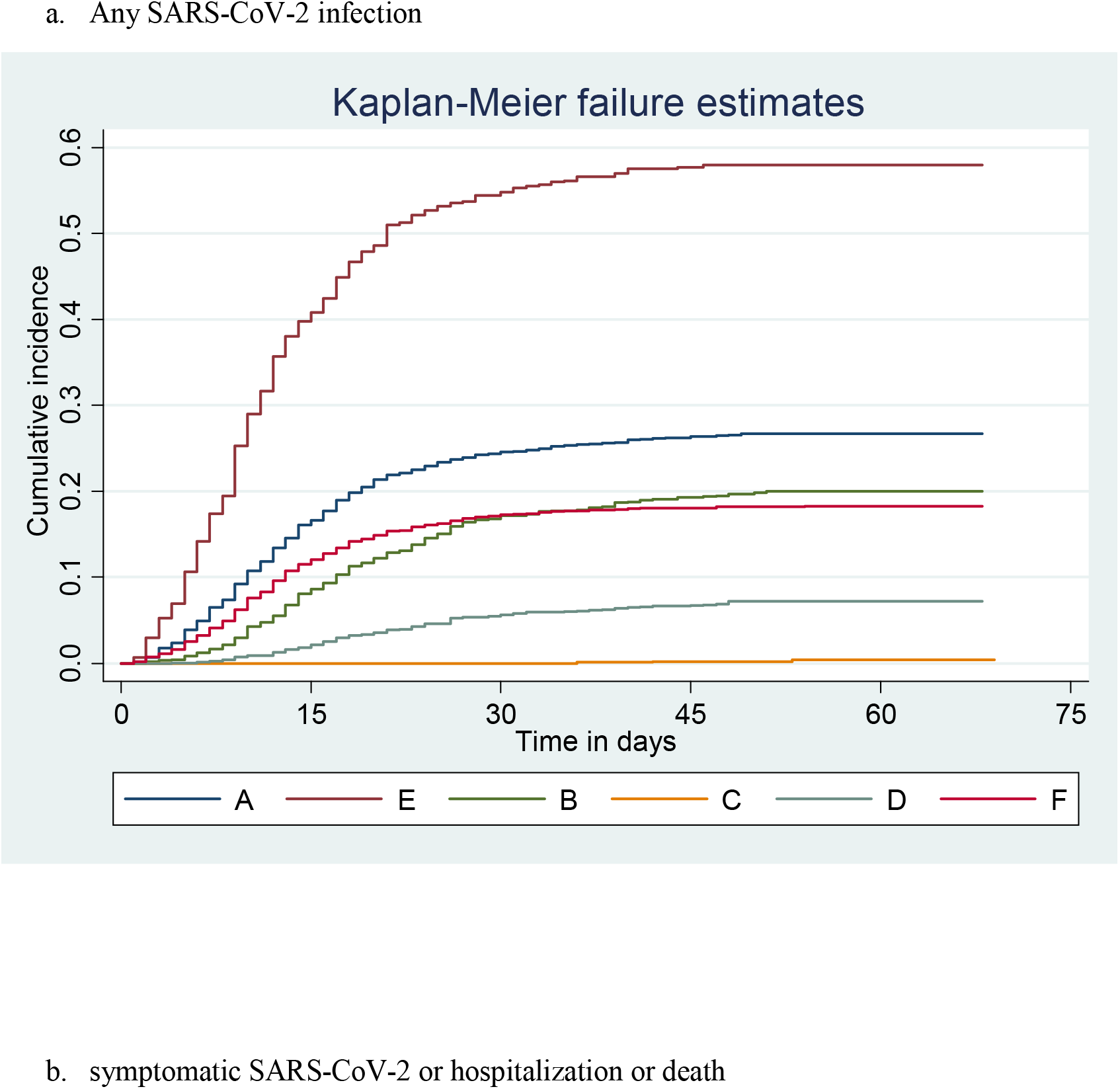
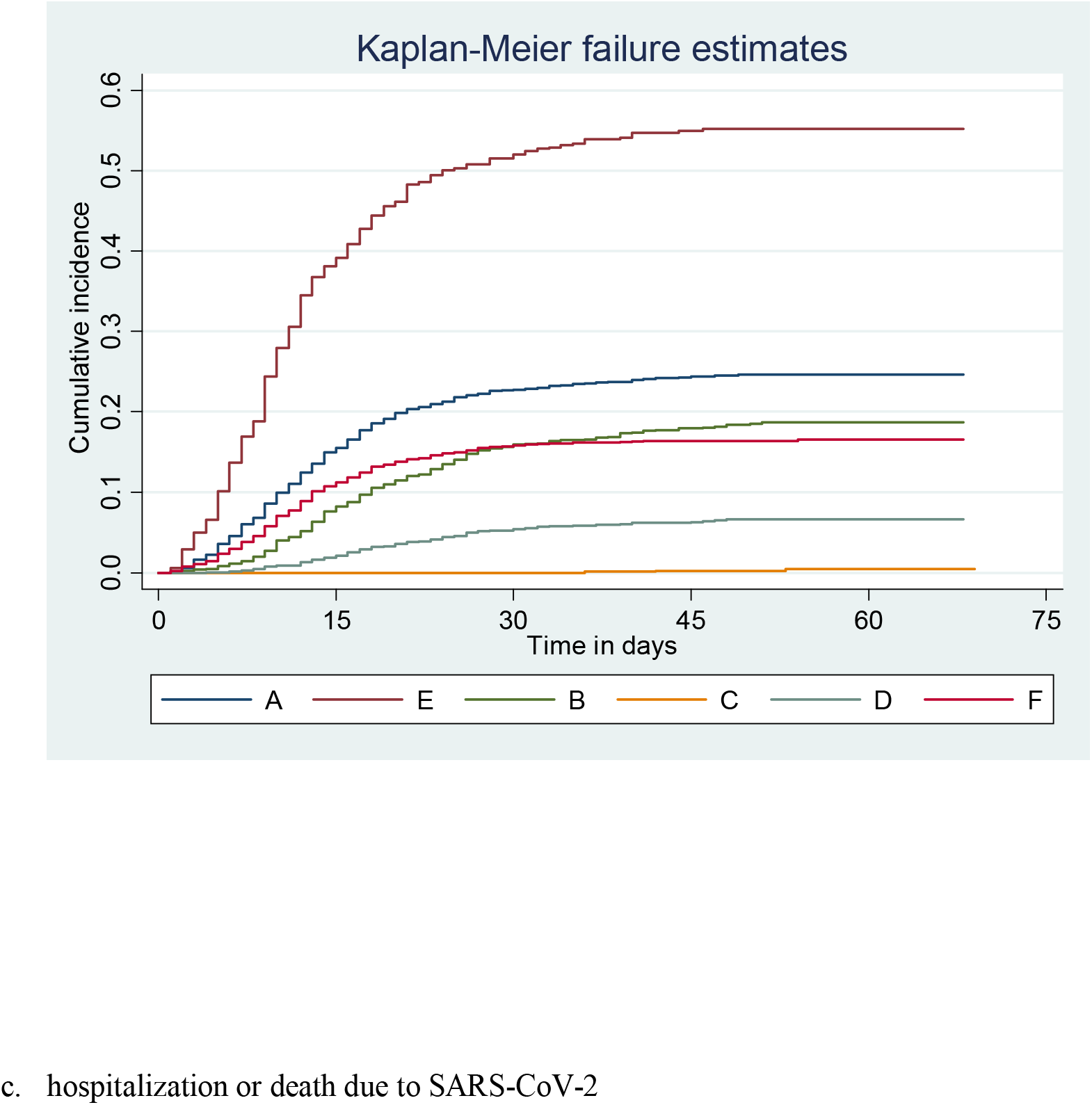
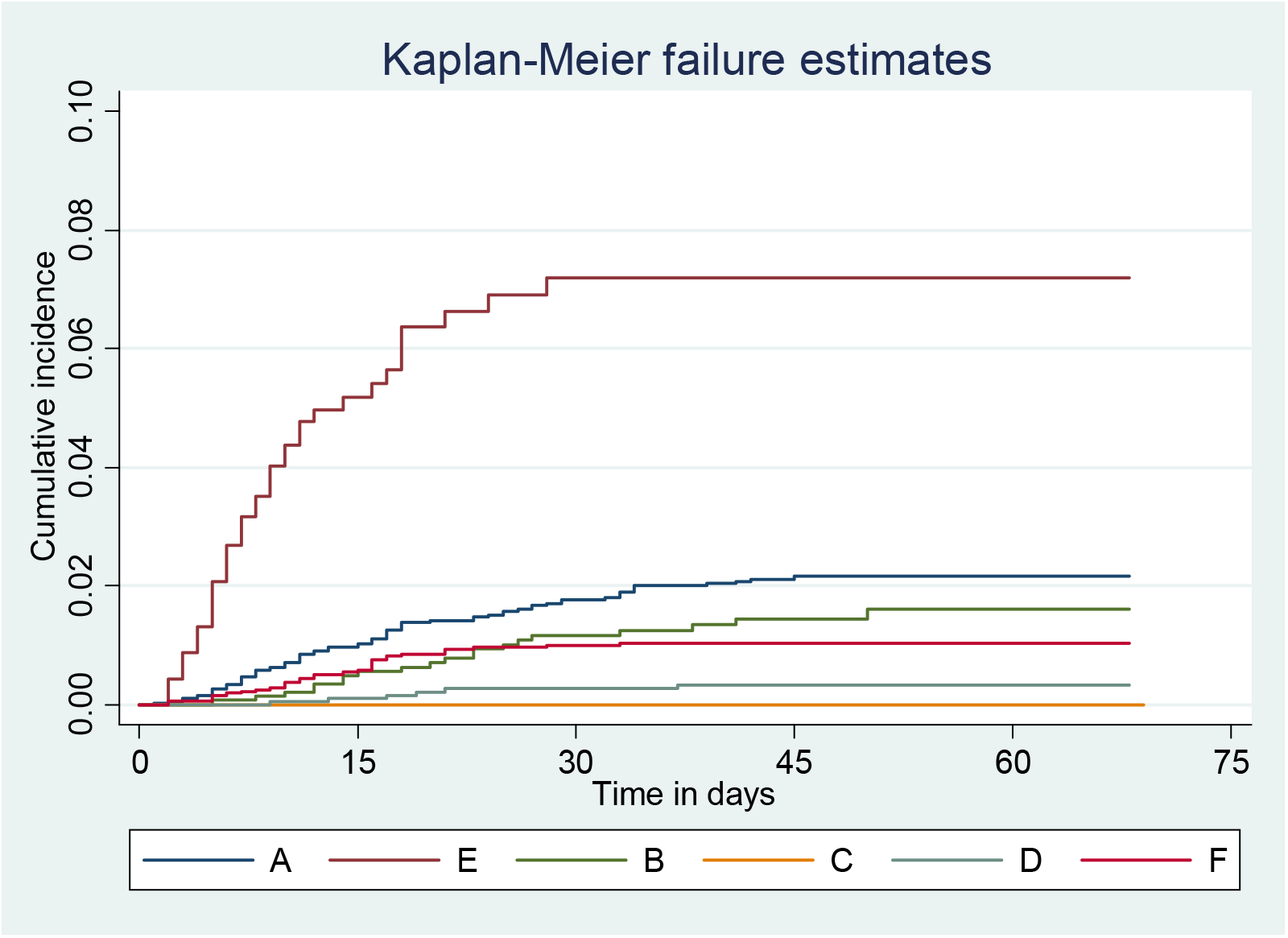
Kaplan-Meier curves showing vaccine effectiveness among the groups for various outcomes a) Any SARS-CoV-2 infection, b) symptomatic SARS-CoV-2 or hospitalization or death, and c) hospitalization or death due to SARS-CoV-2

## Discussion

Our study found modest effectiveness of BBV152 (two doses) as 44% against any SARS CoV-2 infection and 61% against hospitalization or death in real life situation in a background setting of delta variant spread. The Phase 3 trial of BBV152 vaccine reported 65% efficacy against the delta variant (7) and a recent test-negative, case-control study reported 50% (17).

The Vaccine Effectiveness (VE) for BBV152 was found to be lower than other available m-RNA and recombinant vaccines, though higher than reported effectiveness of inactivated Sinovac-CoronaVac vaccine, during resurgent wave in New Delhi, India when delta variant transmission was high. Among HCWs from Brazil against a backdrop of gamma variant transmission, the adjusted VE of CoronaVac 2 doses after two doses was reported to be 37.1% (95% CI -53.3 to 74.2) (20). A study in HCWs in the United States of America reported effectiveness of mRNA-1273 (Moderna) as 96% and BNT162b2 (Pfizer BioNTech) as 89% (21). In a cohort of HCWs based in England, the VE of BNT162b2 was 85%, 7 days after 2 doses (22). The Indian COVID-19 VE estimates have been reported earlier for the Covishield vaccine. In a study from a south Indian multi-speciality hospital, the vaccine protection was reported as 65%, 77%, 92%, and 94% for prevention of infection, hospitalization, need for oxygen therapy and intensive care admission, respectively, though the estimates were unadjusted for any co-variates (14).

The reported estimates vary based on setting, type of vaccine and SARS-CoV-2 variant predominance during data collection period and methods used in estimating effectiveness. VE for m-RNA vaccines and ChAdOx1 nCoV-19 vaccine has been found to be reduced for protection against infection by delta variant (16). A study from the United States (HEROES-RECOVER Cohorts) reported VE during the delta predominant period as 66% (95% CI: 26, 84) compared to 91% (81, 96) in months preceding delta predominance (23).

Convalescent sera of recipients of BBV152 in Indian setting were able to neutralize variants in broader B.1.617 lineage (24). There was a clear contrasting difference between protection offered by single dose, which was low and inconsistent across the entire severity spectrum, compared to two dose schedules. This is corroborated by antibody measurement studies in HCWs that reported only 40% seropositive response after one dose as compared to 80% response after receipt of two doses of BBV152 (25). We found higher vaccine effectiveness response in HCWs who had completed their vaccination after 10 April 2021, indicative of varying effectiveness levels with shorter follow up intervals. On the contrary, the vaccine protection was not found in the partially vaccinated HCWs as on 10 April, indicating other unmeasured variables eliciting the response of the single dose of the vaccine, during the study period. Lower effect of partial dose was also reported with inactivated vaccine CoronaVac, compared to m-RNA vaccines (26). For m-RNA vaccines, such low response for one dose was observed in case of delta variant, corroborated with laboratory neutralization studies (27).

From our institute, a genomic analysis study of 63 vaccine breakthrough infections (that included a sample of health care workers), 53 received BBV152 vaccine, and among them, was the predominant lineage detected in 13 fully vaccinated and 11 partially vaccinated patients (28). Similar data have been reported from other centres (29,30). Breakthrough infections against delta variant have been reported from all the settings around the world against a variety of vaccines, by now (12,13).

Our study had few limitations. In this observational study, some of the confounding due to residual covariates could not be omitted, though we adjusted our VE estimates for the known confounders including age, sex, type of health care worker, body mass index, and comorbidity status. There could be additional covariates like risk averse behaviour adopted by HCWs, that were not measured and reported, but could possibly influence effectiveness estimates. Vaccination status and other data were based on self-reports; however, sufficient details were captured and data were validated to authenticate the exposure and outcome data. We did not perform genomic analysis of SARS CoV-2 infections following vaccination reported in our study, though evidence from other studies was correlated and the delta variant was predominant lineage in transmission during the study period.

We did not collect information about source of exposure of infection in our study HCWs. Also, some of the asymptomatic infections might be missed out, owing to lesser chances of them being tested. We anticipate in our setting that HCWs due to continuous exposures would get themselves tested more due to ready availability of testing and screening facilities within our Institute, though the possibility of varying rates of testing among different categories of HCWs could not be ruled out. HCWs are at increased risk of SARS CoV-2 infection due to occupational exposure and our results apply to them. The estimates for VE might not be generalizable to older people and other general community members. All those who were infected with COVID-19 earlier, prior to the roll-out of the vaccination programme, were excluded in this evaluation. Further, serological and immunological studies will be needed for examining long term immunity offered by the vaccine. Ours is a single site study, thus evidence from more settings will be needed for vaccine effectiveness. The strength of the study lies in generating evidence about BBV152 vaccine in real life situation. We followed STROBE guidelines for reporting the study (31).

## Conclusions

BBV152 vaccine (two doses) offers a modest protection against SARS CoV-2 infection, and symptomatic infection including hospitalization and death, among a cohort of HCWs in real world scenario. Our estimates for VE represent the setting when delta variant was predominant. In the absence of protective response with the partial receipt of BBV152, efforts for maximizing coverage with full dose schedule is vital. It is imperative that HCWs continue with COVID appropriate behaviour with use of personal protective equipment and follow distancing norms in order to maximize public health gains.

## Data Availability

All data produced in the present work are contained in the manuscript

## Acknowledgement

We thank Dr. Rajiv Bahl, Department of Maternal, Newborn, Child and Adolescent Health and Ageing, World Health Organization, Geneva, Switzerland, for providing us scientific advice for analysing the data.

